# Longitudinal brain age prediction and cognitive function after stroke

**DOI:** 10.1101/2022.03.18.22272556

**Authors:** Eva B. Aamodt, Dag Alnæs, Ann-Marie G. de Lange, Stina Aam, Till Schellhorn, Ingvild Saltvedt, Mona K. Beyer, Lars T. Westlye

## Abstract

**Background:** Advanced age and poor brain health have been associated with higher risk for more severe clinical and cognitive outcomes following stroke, but more accurate models for clinical prediction are needed. Machine learning based on brain scans can be used to estimate brain age of individual patients, and the corresponding difference from chronological age, the brain age gap (BAG) has been investigated in a range of clinical conditions. Yet, the predictive value for post stroke NCD has not been established. To this end, using longitudinal data we aimed to investigate the association between BAG and post-stroke neurocognitive disorder (NCD) over time.

**Methods:** 269 stroke survivors (55.4% women, mean (SD) age = 71 (11) years) were included from the Norwegian Cognitive Impairment After Stroke (Nor-COAST) study. MRI and clinical data were collected shortly after the acute stroke (neuropsychological data first after 3 months) and at 18- and 36 months follow-up. The brain age model was trained in an independent sample using established protocols based on machine learning and brain structural features from Freesurfer and applied to the current dataset. We used linear mixed effects (LME) models and survival analysis to assess the associations between cognitive status and longitudinal brain age.

**Results:** LMEs revealed a main effect of BAG on post stroke NCD across time, confirming our main hypothesis that a younger appearing brain may protect against post stroke NCD. For patients with no NCD at baseline, survival analysis suggested that higher baseline BAG was associated with higher risk of post stroke NCD at 18 and 36 months.

**Conclusion:** A younger appearing brain is associated with lower risk of post stroke NCD up to 36 months after a stroke, even among those showing no evidence of impairments 3 months after hospital admission. Brain age prediction based on brain scans provides a reliable assessment of brain structure and is sensitive to post stroke NCD with predictive value for cognitive decline among patients with no impairment at the initial assessment.

## 1. Introduction

Stroke increases risk of functional and cognitive decline, and 35-50% of stroke survivors fulfil criteria for mild or major neurocognitive disorder (NCD, previously ‘mild cognitive impairment’ and ‘dementia’, respectively) (American Psychiatric Association, 2013) 15 months after a mild stroke (Jacquin et al., 2014; Weaver et al., 2021). While cognitive impairments are prevalent among stroke survivors, a substantial proportion remains largely unaffected or experience improvements or complete remission after initial impairments during the acute phase (Levine et al., 2015). It is well known that poor health and incipient neurocognitive deficits prior to the stroke is associated with poor outcome. However, less is known about resilience factors. The high prevalence and burden of post-stroke cognitive impairments warrant further studies to identify malleable protective factors, with implications for public health policies and prevention strategies as well as post stroke treatment and rehabilitation approaches to reduce the burden of post stroke cognitive impairments.

Several factors are associated with poorer outcomes after stroke, including old age, lower education, pre-stroke disability, left-sided stroke (Pendlebury, 2012; Pendlebury & Rothwell, 2009), diabetes mellitus, a history of stroke (Lo et al., 2019), larger stroke lesions and vascular factors, such as white matter hyperintensities (WMH) (Schellhorn et al., 2021a; Aamodt et al., 2021). Brain atrophy after stroke (Brodtmann et al., 2020; Levine et al., 2015) is also associated with cognitive decline (Mijajlovic et al., 2017; Haque et al., 2019), particularly if accompanied by hypertension (Sayed et al., 2020) or other vasculopathies (Schellhorn et al., 2021b), with global and medial temporal lobe atrophy and WMH load among the most predictive factors 12 months post-stroke (Jokinen et al., 2020; Casolla, 2019; Wang et al., 2021; Ball et al., 2021; Schellhorn et al., 2021a).

Advanced age is a major risk factor for both stroke and of poorer prognosis following stroke (Pendlebury & Rothwell, 2009), and is the primary risk factor for neurodegeneration (Hou et al., 2019) and lower cognitive reserve and resilience, rendering the individual more vulnerable to post stroke NCD (Mattson & Arumugam, 2018;Hou et al., 2019; Aamodt et al., 2021). The individual differences in cognitive function among seniors are substantial (Sanchez-Izquierdo & Fernandez-Ballesteros, 2021) and chronological age alone is therefore not a reliable predictor for pre-stroke function and post stroke outcome (Wagner et al., 2016). Sensitive biomarkers or tools for assessing and monitoring biological age and aging may have great value for predicting decline and resilience following stroke. Current advances have allowed for an accurate estimation of an individual’s brain age based on machine learning on structural brain features from clinically accessible brain scans (Cole & Franke, 2017; Cole et al., 2017). Discrepancy between the predicted and chronological age, also termed the brain age gap (BAG; with lower values indicating a younger appearing brain) has been used as an intuitive, reliable and sensitive surrogate marker for brain health, various brain disorders, and cognitive aging (Wrigglesworth et al., 2021; Boyle et al., 2021; Kaufmann et al., 2019), and for predicting clinical conversion from mild to major NCD (Gaser et al., 2013).

With some exceptions (Richard et al., 2020) few brain age studies have been conducted on stroke patients and post stroke NCD (Wrigglesworth et al., 2021), and a careful evaluation of the clinical and predictive value of brain age among stroke survivors is warranted. To this end, using longitudinal brain imaging at baseline, 18- and 36 months, and clinical data obtained at baseline, 3-, 18- and 36 months post stroke we tested the following hypotheses: I) patients with no or minimal post stroke cognitive impairments exhibit lower BAG than patients with post stroke cognitive impairments, II) lower BAG at baseline is associated with less cognitive impairments at 18 and 36-months follow-up among patients showing normal cognitive function at baseline, and III) patients showing preserved cognitive function across the follow-up period show less increase in BAG over time compared to patients showing evidence of cognitive decline.

## 2. Material and methods

### 2.1 Sample and inclusion criteria

The study is based on data from the Norwegian Cognitive Impairment After Stroke study (Nor-COAST) - a prospective longitudinal multicenter cohort study recruiting patients hospitalized with acute stroke at five Norwegian stroke units (Thingstad et al., 2018). Patient recruitment started in May 2015 and was completed in March 2017. Study details are described elsewhere (Thingstad et al., 2018). Nor-COAST was approved by the regional committee for medical and health research, REK Nord (REK number: 2015/171), and registered on clinicaltrials.gov (NCT02650531). REK Nord has also approved this current sub study (REK number: 2019/397). All participants provided written informed consent in accordance with the Declaration of Helsinki. If a participant was unable to give consent, written informed consent for participation was provided by a family proxy.

Inclusion criteria for Nor-COAST: (a) patients admitted with acute ischemic or hemorrhagic stroke hospitalized within one week after onset of symptoms, diagnosed according to the World Health Organization (WHO) criteria; (b) age over 18 years; (c) fluent in a Scandinavian language. Exclusion criteria for Nor-COAST: (a) not treated in the participating stroke units; (b) symptoms explained by other disorders than ischemic brain infarcts or intracerebral hemorrhages; (c) expected survival less than three months after stroke. Additional inclusion criteria for the MRI study: (a) modified Rankin scale (mRS) <5 before the stroke; (b) able and willing to perform MRI. Reasons for declining participation in the MRI sub study were not recorded, in line with ethical guidelines.

### 2.2 Clinical characteristics

Demographic and clinical data were collected by study nurses and stroke physicians. Based on previous literature, the following baseline factors were included; age, years of education, sex, waist-to-hip ratio (WHR), stroke severity measured with the National Institute of Health Stroke Scale (NIHSS), disability measured using mRS, atrial fibrillation (AF), diabetes mellitus, and pre-stroke hypertension and global cognition, based on the global deterioration scale (GDS, Reisberg et al., 1982). NIHSS ranges from 0 to 42, with higher scores indicating more severe strokes. mRS ranges from 0 to 6, with higher scores indicating worse disability (Wilson et al., 2005). As described in Munthe-Kaas et al. (2020), AF was defined as a history of permanent or paroxysmal AF or atrial flutter detected in electrocardiogram and described in medical records and/or permanent or paroxysmal AF or atrial flutter detected in electrocardiogram and/or telemetry during hospital stay. Diabetes mellitus was defined as a history of diabetes mellitus from medical records and/or pre-stroke use of antidiabetic medication and/or HbA1c≥6.5% at admittance for stroke. Hypertension was defined as pre-stroke use of antihypertensive medication. GDS ranges from 1 to 7 and pre-stroke GDS was collected through interviews with relatives or caregivers.

### 2.3 Neurocognitive and neuropsychiatric assessment

The neuropsychological test battery included Trail making A and B (TMT A and B) (Reitan, 1958), ten word memory and recall test (CERAD) (Morris et al., 1988), the controlled oral word association test (COWAT) (Loonstra, Tarlow & Sellers, 2001), the Montreal Cognitive Assessment (MoCA) (Nasreddine et al., 2005), the Ascertain Dementia 8-item informant questionnaire (AD-8) (Galvin et al., 2005), and the Global Deterioration Scale (GDS) (Reisberg et al., 1982).

Criteria for NCD were based on the Diagnostic and Statistical Manual of Mental Disorders (DSM-5), encompassing neuropsychological assessment and instrumental activities of daily living (I-ADL) <u>(American Psychiatric Association, 2013)</u>. Patients scoring >1.5 SD below the normative mean on at least one of the cognitive domains tested (attention, executive function, learning and memory, language, or perceptual-motor function) were defined as having post stroke NCD. Major NCD was defined as post stroke NCD accompanied by dependency in I-ADL, whereas mild NCD was defined as post stroke NCD with independencies in any I-ADL (Munthe-Kaas et al., 2020). NCD status was grouped into no NCD and any NCD (mild NCD plus major NCD).

### 2.4 MRI acquisition and processing

MRI was performed at baseline (within 2-7 days of the acute stroke), and at 18-and 36 months follow-up. The assessments were done at five hospitals (supplementary table 4), using a single scanner at each site (GE Discovery MR750, 3T; Siemens Biograph_mMR, 3T; Philips Achieva dStream, 1.5T; Philips Achieva, 1.5T; Siemens Prisma, 3T). The MRI protocol comprised 3D-T1 weighted, axial T2, 3D-Fluid attenuated inversion recovery (FLAIR), diffusion-weighted imaging (DWI), and susceptibility-weighted imaging (SWI) sequences (details in supplementary table 3).

### 2.5 Stroke volume extraction and stroke location

As previously described (Schellhorn et al., 2021a), lesion masks were generated based on the DWI data obtained in the acute/-sub-acute phase, and were semi-automatically labeled and quantified using ITK-Snap snake tool (v. 3.8.0) (Yushkevich et al., 2006). Stroke location was determined using the Talairach lobe atlas (Lancaster et al., 1997; Lancaster et al., 2000). The labels ‘anterior lobe’ and ‘posterior lobe’ were then gathered into ‘cerebellum’, and ‘medulla’, ‘midbrain’, and ‘pons’ were gathered into ‘brainstem’. If the stroke was labeled as ‘background’, the Harvard-Oxford structural atlas (Makris et al., 2006; Frazier et al., 2005; Desikan et al., 2006; Goldstein et al., 2007) was used. Stroke location was established as the location with the highest probability, as set by the atlas. For participants with multiple lesions, the location of the largest lesion was used.

### 2.6 WMH segmentation

As previously described (Aamodt et al., 2021), WMH segmentation was performed using the fully automated and supervised tool BIANCA (Griffanti et al., 2016). BIANCA is based on an algorithm using the k-nearest neighbor and classifies the probability of WMHs based on intensity and spatial features of voxels. The intensity- and spatial based lesion classification probability threshold was set to 0.7. WMH volume was calculated in mm^3^ and converted to millilitre (ml).

### 2.7 Brain age prediction

Features used for brain-age prediction were generated through cortical reconstruction and parcellation, and volumetric segmentation of the 3D-T1 scans using the longitudinal stream of Freesurfer 6.0.1 (http://surfer.nmr.mgh.harvard.edu/) (Fischl, 2012), including brain extraction, intensity normalization, tissue segmentation and surface reconstruction. Quality control of the FreeSurfer output was conducted based on the ENIGMA protocol (http://enigma.ini.usc.edu) supplemented by Euler numbers extracted from FreeSurfer (Rosen et al., 2018). Scans were discarded if involving major errors in reconstruction, segmentation, or parcellation, or involving extensive motion or other artefacts.

We used machine learning to predict the age of each patient based on cortical thickness, surface area, and gray-and white matter volume. This approach has revealed highly accurate and reliable estimates in stroke patients (Richard et al., 2019).

For model training we used an independent sample of healthy controls (n = 934, age range = 18-88 years, mean = 55.8, SD = 17.4) from the Cambridge centre for Aging and Neuroscience (CamCAN) (Shafto et al., 2014; Taylor et al., 2017) and StrokeMRI (Richard et al., 2018; Sanders et al., 2021). In line with previous work (Kaufmann et al., 2019; Richard et al., 2020; Høgestøl et al., 2019; Beck et al., 2022a; Anatürk et al., 2020), the age-prediction model was trained using XGBoost regression (Chen & Guestrin, 2016) in Python 3 with scikit-learn (Pedregosa et al., 2011) including nested cross-validation for hyperparameter tuning and model evaluation (5 inner and 10 outer folds). The prediction model was run using both global and regional feature inputs, and based on the Desikan-Killiany atlas (Desikan et al., 2006) and subcortical segmentation.

Next, we applied the trained model on previously unseen data from the Nor-COAST patients. To quantify model performance, we calculated the Pearson’s correlation between predicted and chronological age and mean absolute error (MAE) in the test set. BAG was computed per scan per individual, and defined as the difference between estimated and chronological age (Figure 1 for examples). To account for age-related bias in the age prediction (Le et al., 2018; de Lange & Cole, 2020), a linear model was used to regress out the main effect of age, age^2^, and sex.

**Figure 1:**
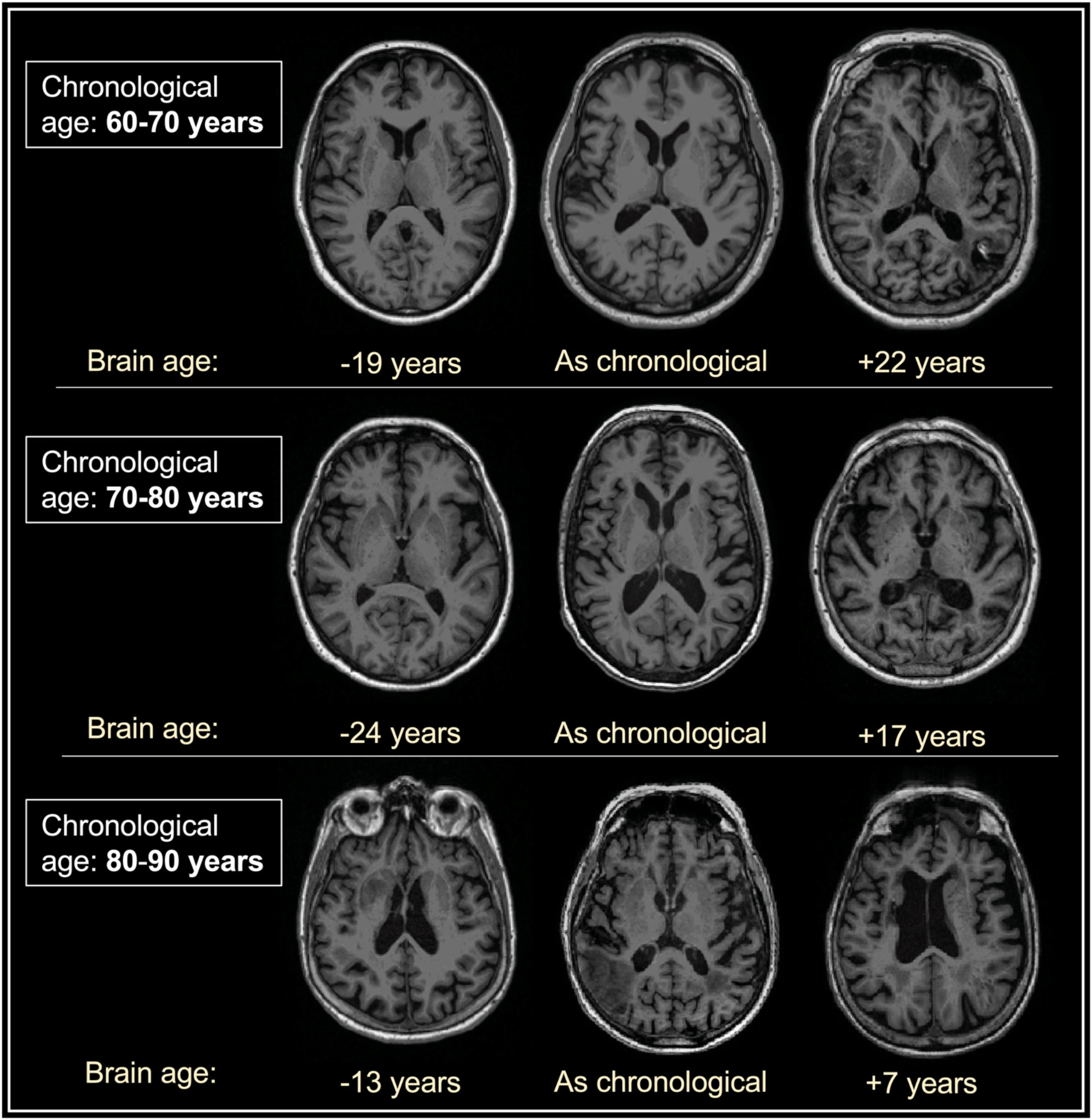
Examples of groups of three patients with same exact chronological age and differing brain age. *Chronological age has been scrambled in order to preserve privacy.

### 2.8 Statistical analysis

Statistical analyses were performed using R v. 3.3.3 (R Core Team 2017) and Stata v. 16. Means and standard deviations (SD) were calculated and outliers and normality checked using histograms. The three main hypotheses outlined above were tested using the following approaches:

First, to test if patients with minimal or no post stroke cognitive impairments exhibited lower BAG than patients with cognitive impairments, we performed one linear mixed effects (LME) model with BAG as dependent variable, patient as random variable and NCD status (no NCD vs. any NCD), timepoint and age as independent variables. An additional model including the interaction between NCD status and time point was also tested. To complement the LME model, we performed three student t-tests comparing BAG between the two NCD groups at each time point, and a survival analysis with NCD status (any NCD) as outcome stratified by BAG (quartiles for cognitive impairments, at 18- and 36-months follow-up).

Next, among patients with normal cognitive function at baseline we performed one LME model with BAG as dependent variable, patient as random variable and NCD status (dichotomous), timepoint and age as independent variables. An additional model including the interaction between NCD status and time point was also performed. In addition, we performed two student t-tests to compare BAG between the NCD groups at 18 and 36 months and a survival analysis with NCD status as outcome stratified by BAG (quartiles) at baseline.

Lastly, to test if patients with preserved cognitive function across the three-year follow-up period show smaller increases in BAG over time compared to patients with cognitive decline, we performed one LME with the dependent variable of BAG, patient as random variable and NCD status change (dichotomous, preserved vs. declined), time point and age as independent variables. An additional model including the interaction between NCD status and time point was also performed. To complement the LME model, we performed two student t-tests to compare BAG between groups (preserved vs. declined) at 18- and 36 months and a survival analysis with NCD status change as outcome stratified by BAG (quartiles) at baseline.

Normality of residuals from the LME models were assessed through visual inspection of quantile-quantile-plots. Margins were calculated and margin plots and survival plots were created. Significance threshold was set to p<.05 after corrections for multiple comparisons. In addition, we performed several analyses to identify variables associated with attrition (supplementary table 6).

## 3. Results

### 3.1 Study population

Figure 2 summarizes the study outline and population. From the complete Nor-COAST population, 352 (43.2%) patients performed baseline MRI which passed quality assessment. Of the MRI group, mean (SD) age at baseline was 72.8 (11.2) years, 157 (44.6%) were women, and average NIHSS score was 4 (4.9). NCD status was missing for 45 patients and Freesurfer reconstruction failed for 38 (10.8%), resulting in a final sample of 269 (76.4% of the total MRI sample) at baseline (55.4% women, mean age = 71 (11) years, education = 12.4 (3.6) years, NIHSS = 3.6 (4.5)). At the neuropsychological testing at three months, 79 (29.4%) and 61 (22.7%) patients fulfilled criteria for mild or major NCD, respectively, resulting in a group of 140 (52%) patients with any NCD and 129 (48%) with no evidence of NCD at three months.

**Figure 2:**
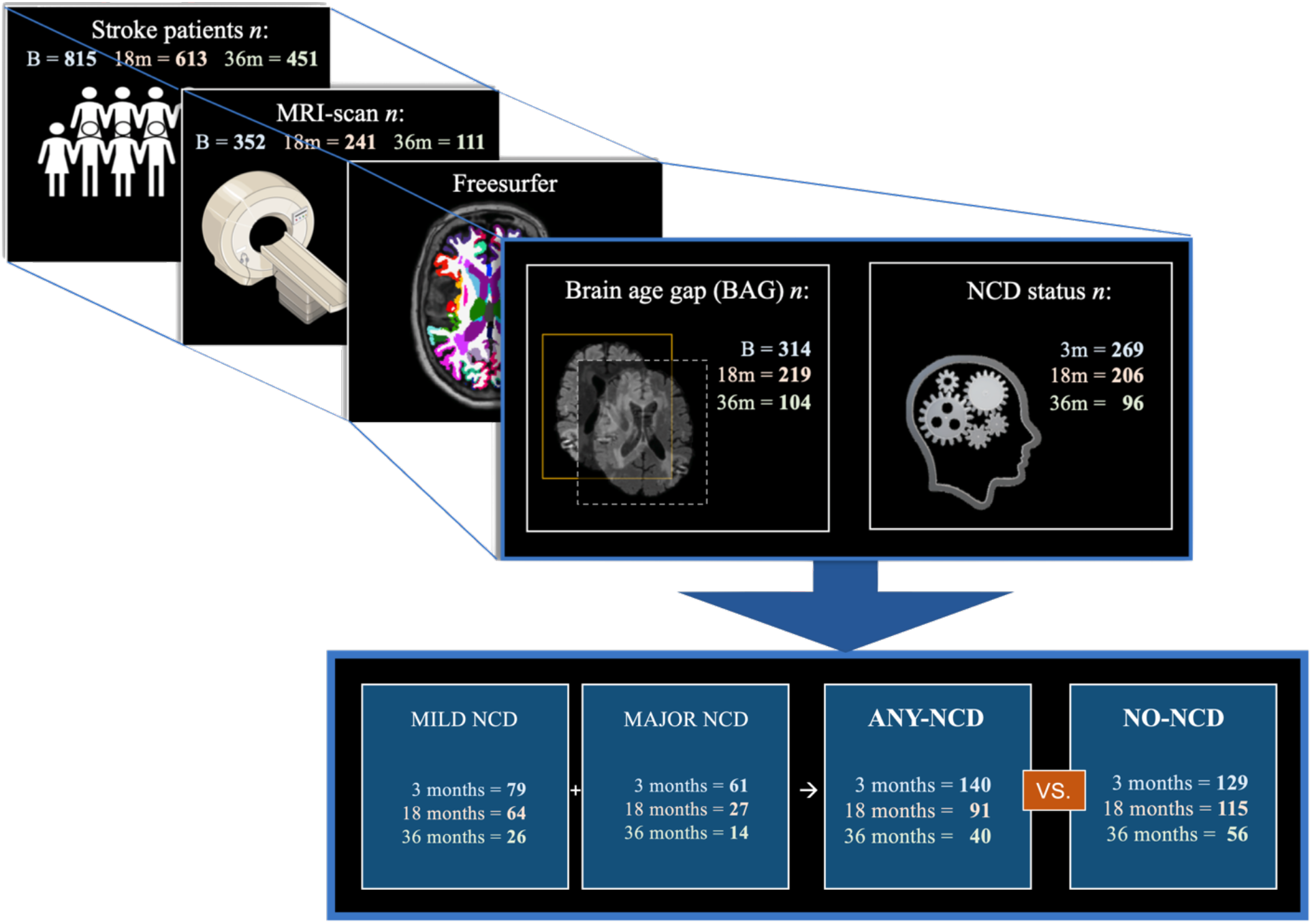
Study population selection and NCD-group categorization by all three timepoints (B (baseline), 18-(18m) and 36 months (36m)). Ending in the two study groups of Any-NCD (mild NCD + Major NCD) and No-NCD. *Baseline NCD status collected at 3 months post-stroke.

### 3.2 Baseline clinical and stroke characteristics

Table 1 summarizes demographic and clinical patient characteristics. At baseline, 167 (69.3%) patients were overweight (WHR>.89 for men and WHR>.98 for women (WHO, 2008)). Other baseline comorbidities and risk factors included hypertension (n = 137, 50.1%), AF (n = 55, 20.4%), and diabetes type 1 or type 2 (n = 50, 18.6%). Pre-stroke cognitive decline (GDS score of 3 to 7) was found in 21 (7.4%) patients (*M* = 1.4 *SD* = .73, min = 1, max = 5). Mean NIHSS scores of 3.58 (4.51, min = 0, max = 42) indicated mainly mild strokes.

**Table 1:** Table depicting demographic, clinical, stroke characteristics and BAG for baseline, 18 months and 36 months, for those with no-NCD, any-NCD and all participants. BAG = Brain age gap. NIHSS = National Institutes of Health Stroke Scale. AF = Atrial fibrillation. GDS = Global Deterioration Scale. MoCA = Montreal Cognitive Assessment. WMH = White matter hyperintensities. * = Missing data. ^b^ baseline. ^C^ Overweight defined as waist-hip-ratio (WHR) = >.86 in women and >1.00 in men. ^D^ BAG defined as chronological age subtracted from brain age.

|  | No-NCD |  |  | Any-NCD |  |  | All |  |  |
| --- | --- | --- | --- | --- | --- | --- | --- | --- | --- |
|  | Baseline | 18 mo. | 36 mo. | Baseline | 18 mo. | 36 mo. | Baseline | 18 mo. | 36 mo. |
| *N* | **129** | **115** | **56** | **140** | **91** | **40** | **269** | **206** | **96** |
| Age | 68.9<br>(11.5) | 69.5<br>(12.4) | 72.7 (8.8) | 72.9<br>(10.3) | 74.2<br>(8.7) | 78.7 (8.3) | 71.0<br>(11.0) | 71.6<br>(11.2) | 75.2 (9.1) |
| Women | 69<br>(53.5%) | 66<br>(57.4%) | 31<br>(55.4%) | 80<br>(57.1%) | 52<br>(57.1%) | 26 (65%) | 149 (55.4) | 118<br>(57.3%) | 57<br>(59.4%) |
| Education<br>(yrs) | 13.2 (3.8) | 13.5<br>(4.1) | 13.4 (4.0) | 11.8 (3.4) | 12.0<br>(3.3) | 11.6 (3.6) | 12.4 (3.6) | 12.8 (3.9) | 12.7 (3.9) |
| Over-weight<sup>b,c</sup> | 77 (67%) | 75<br>(74.3%) | 12<br>(66.7%) | 90<br>(71.4%) | 58<br>(72.5%) | 9<br>(64.3%) | 167<br>(69.3%) | 133<br>(73.5%) | 21<br>(65.6%) |
| NIHSS<sup>b</sup> | 3.2 (3.8) | 4.0 (5.8) | 4.0 (6.7) | 3.9 (5.0) | 3.3 (3.6) | 4.2 (4.1) | 3.6 (4.5) | 3.7 (3.9) | 4.1 (5.7) |
| AF<sup>b</sup> | 21<br>(16.3%) | 22<br>(19.1%) | 10<br>(17.9%) | 34<br>(24.3%) | 27<br>(29.7%) | 10 (25%) | 55<br>(20.4%) | 49<br>(23.8%) | 20<br>(20.8%) |
| Diabetes<sup>b</sup> | 20<br>(15.5%) | 22<br>(19.1%) | 9<br>(16.1%) | 30<br>(21.4%) | 17<br>(18.7%) | 6 (15%) | 50<br>(18.6%) | 39<br>(18.9%) | 15<br>(15.6%) |
| Hyper-<br>tension<sup>b</sup> | 57<br>(44.2%) | 56<br>(48.7%) | 26<br>(46.4%) | 80<br>(57.1%) | 52<br>(57.1%) | 25<br>(62.5%) | 137<br>(50.1%) | 108<br>(52.4%) | 51<br>(53.1%) |
| mRS* | 1.73<br>(1.78) | .97 (.75) | 1.26<br>(1.26) | 2.23<br>(1.23) | 1.57<br>(1.10) | 1.92<br>(1.55) | 1.99<br>(1.23) | 1.24 (.97) | 1.58 (1.43) |
| GDS* (pre-stroke) | 1.16 (.39) | 1.13<br>(.34) | 1.12 (.31) | 1.55 (.91) | 1.37<br>(.72) | 1.38 (.67) | 1.36 (.73) | 1.24 (.56) | 1.22 (.51) |
| MoCA* | 26.3 (2.7) | 26.6<br>(2.9) | 20.4<br>(10.5) | 22.0 (5.0) | 22.8<br>(4.9) | 22.3 (7.3) | 24.1 (4.6) | 24.9 (4.3) | 21.2 (9.3) |
| BAG<sup>d</sup> | -1.91<br>(8.16) | -1.83<br>(8.99) | -3.36<br>(9.16) | 2.11<br>(9.38) | .84<br>(9.38) | .92 (9.94) | .18 (9.03) | -.65<br>(9.24) | -1.58<br>(9.67) |
| Stroke<br>volume<br>(ml)<sup>*b</sup> | 7.23<br>(11.03) | 5.41<br>(9.45) | 5.62<br>(9.23) | 10.33<br>(21.77) | 11.04<br>(21.06) | 12.88<br>(27.03) | 8.90<br>(17.67) | 7.96<br>(15.96) | 8.41<br>(18.40) |
| WMH<br>(ml)<sup>*b</sup> | 16.74<br>(14.78) | 16.18<br>(13.20) | 15.94<br>(14.29) | 27.62<br>(25.20) | 23.17<br>(24.19) | 19.13<br>(18.94) | 22.47<br>(4.59) | 19.41<br>(19.31) | 18.18<br>(16.19) |

Mean (SD) stroke lesion volume was 8.6 (17.4) ml. Right hemisphere lesions were most frequent (51.4%), and 6 (2.9%) patients showed acute bilateral strokes. The most frequent stroke locations were the frontal lobe (28.8%) and subcortical structures (20.6%, supplementary table 5).

The NCD group had higher risk scores compared to the no NCD group, with higher age (72.9 (10.3) vs. 68.9 (11.5)), more women (57.1% vs. 53.5%), lower baseline MoCA scores (22.0 (5.0) vs. 26.3 (2.7)), higher baseline BAG (2.11 (9.38) vs. -1.91 (8.16)), larger stroke volume (10.33 (21.77) vs. 7.23 (11.03)), and larger WMH volume (27.62 (25.20) vs. 16.74 (14.78)).

### 3.3. BAG and NCD across time

Figure 3 shows the results from the LMEs and survival analyses. LMEs revealed a significant main effect of NCD status on BAG (coef. = 1.37, *p =* .010), and no significant effect of time (coef. = -.30, *p* = .23). We found no significant interaction between NCD status and time (coef. = .50, *p* = .33). Further summary statistics are reported in supplementary table 1 and 2. T-tests revealed higher BAG in the NCD group (*M* = 2.11, *SD* = .79, *n =* 140) than the no NCD group (*M* = -1.91, *SD* = .72, *n =* 129) at baseline (*t*(267) = -3.73, *p*<.001). At 18 months, BAG was higher among patients with NCD (*M* = .84, *SD* = .72, *n =* 91) than those without NCD (*M* = -.1.83, *SD* = .84, *n =* 115) (*t*(204) = -2.08, *p =* .04). Similar patterns were found at 36 months, with higher BAG in the NCD group (*M* = .92, *SD* = 1.57, *n =* 40) than the no NCD group (*M* = -.3.36, *SD* = 1.22, *n =* 56) (*t*(94) = -2.18, *p =* .03).

**Figure 3:**
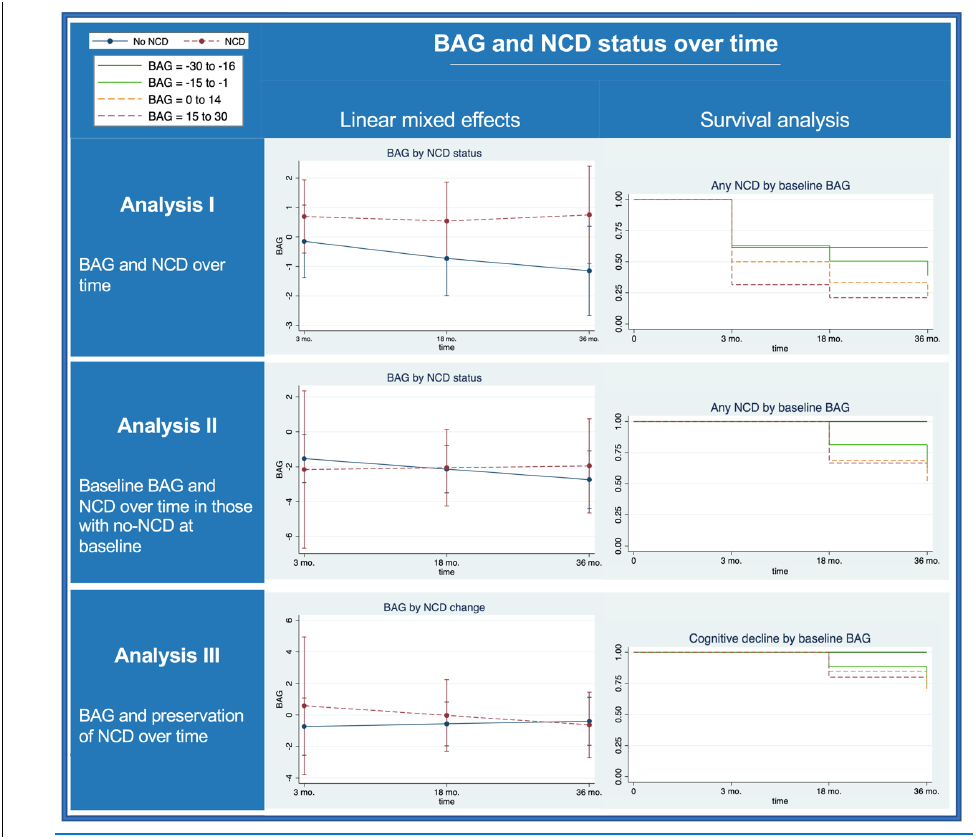
Plots of predicted margins for BAG by NCD status (or change), and Kaplan-Meier estimation plots of survival analysis of any-NCD (or cognitive decline) by baseline BAG, per analysis. BAG = Brain age gap. NCD = neurocognitive disorder. Mo = months.

Survival analysis revealed a significant difference between the four BAG groups in having any NCD over time (*p =* .01), such that patients in the upper BAG quartile at baseline (+15 to +33 yrs.) had a higher chance of having NCD over time.

### 3.4 Baseline BAG and NCD over time in those with no-NCD at baseline

Among the 89 patients with no NCD at baseline LMEs revealed no significant main effect of NCD status on BAG (coef. = .56, *p =* .50) and no significant effect of time (coef. = -.57, *p =* .095). We found no significant interaction between NCD status and time (coef. = .71, *p =* .65). T-tests revealed no significant BAG differences at baseline between patients with (*M* = .51, *SD* = 1.69, *n =* 22) and without (*M* = -.2.47, *SD* = 1.03, *n =* 67) NCD at 18 months (*t*(87) = -1.46, *p =* .15). Similarly, no significant group difference in BAG was found between the NCD (*M* = .42, *SD* = 2.74, *n =* 10) and the no NCD (*M* = -1.34, *SD* = 1.49, *n =* 37) group at 36 months (*t*(45) = -.55, *p =* .59). Survival analysis showed a significant difference between the four BAG groups in having NCD over time for those with no NCD at baseline (*p =* .008), such that the groups with the upper BAG quartiles (0 to +14 and +15 to +30) had a higher risk of NCD at 18- or 36 months.

### 3.5 BAG and preservation of NCD over time

LMEs revealed no significant main effect of cognitive decline on BAG (coef.= .27, *p* = .71) and no significant effect of time (coef. = .04, *p =* .92). We found no significant interaction between NCD status change and time (coef. = -.78, *p =* .60). T-tests revealed no significant group differences in BAG at 18 months between preserved NCD status (*M* = -.70, *SD* = .80, *n =* 142) or cognitive decline (*M* = .63, *SD* = 1.66, *n =* 26) from baseline to 18 months (*t*(166) = -.066, *p =* .51). No significant group difference was found of BAG at 36 months between preserved NCD status (*M* = -1.86, *SD* = 1.31, *n =* 59) or cognitive decline (*M* = .86, *SD* = 2.04, *n =* 17) from baseline (*t*(74) = -1.01, *p =* .31). Survival analysis showed no significant difference between the four BAG groups in having cognitive decline (as opposed to preserved NCD) over time (*p =* .49).

## 4. Discussion

By the use of longitudinal brain age prediction and neurocognitive assessment in a clinical context we tested the hypotheses that I) brain structural characteristics are associated with post stroke cognitive function and impairments, II) a younger appearing brain among patients showing normal cognitive function at baseline is associated with lower risk of cognitive impairments at 18 and 36- months follow-up, and III) patients showing preserved cognitive function from baseline and across the follow-up period show less evidence of brain aging over time compared to patients showing cognitive decline. Our analyses revealed that higher BAG was associated with post stroke cognitive impairment across time, confirming our main hypothesis that a younger appearing brain may reflect relevant protective factors against cognitive decline after stroke. Next, survival analysis of patients with no NCD at baseline suggested that higher BAG at baseline was associated with NCD at 18 and 36 months. Lastly, our findings revealed no significant difference in BAG between the group showing preserved versus the group showing a declined cognitive function from baseline and across follow-up times.

### 4.1 Brain structural characteristics associated with post stroke cognitive function and impairments

In line with our hypothesis, longitudinal analysis across all patients and assessments revealed a significant association between brain age and cognitive impairment across all time points, indicating more impairment among patients with higher BAG. A previous longitudinal study reported reliable brain age prediction among chronic stroke patients, and, while patients with lower BAG showed marginally better cognitive performance, no significant associations with post stroke cognitive function were found (Richard et al., 2020). A likely explanation for the diverging results is that the previous study included relatively young and mild cases with less cognitive impairment than the current study. Also, the current sample is larger, which offers more statistical power.

Advanced age is a risk factor for stroke and post stroke cognitive impairment, and younger age is associated with better prognosis following stroke (Pendlebury & Rothwell, 2009; Mattson & Arumugam, 2018). While the current study population had a relatively high mean age, several patients displayed both a younger appearing brain and lack of post stroke cognitive impairment. This is not too surprising, as the strokes in this cohort were overall relatively small. However, this finding is also in line with the idea that brain age is a better predictor of post stroke cognitive outcome than what chronological age is.

Aging is also associated with an overall vulnerability to both peripheral and central nervous system disease, likely partly due to an overall decreased maintenance and damage repair on cell level (Rattan, 2012) and the possible increased burden of multi-morbidity (Bektas et al., 2018). Around half of the participants in the current study suffered from pre-stroke hypertension, and a previous study on the same cohort has shown high levels of pre-stroke brain pathology (Schellhorn et al., 2021b). These diseases render the brain at risk and more vulnerable to the consequence of future events (Kelly et al., 2021). One example of this is through systemic inflammation, which has been found to cause neurodegeneration (Marogianni et al., 2020; Yang et al., 2020). Another example is cerebral small vessel disease, often caused by hypertension (Paolini Paoletti et al., 2021), which leads to brain tissue atrophy, but also an enhanced risk of NCD on its own. A stroke may thereby exacerbate on two different levels; compromising or destroying brain tissue in an already atrophying brain, and by introducing a taxing event on an already taxed system. An ischemic stroke leads to inflammatory responses both acutely and long-term (Shi et al., 2019; Low et al., 2019). Chronic inflammation will aggravate existing diseases but also factors associated with aging (Germolec et al., 2018) and cognitive decline (Darweesh et al., 2018; Gabin et al., 2018; Zheng & Xie, 2018). Pre-existing diseases (Cipolla et al., 2018) and brain pathology are common in stroke patients, in particular among those who develop cognitive impairment (Schellhorn et al., 2021b).

### 4.2 Brain age and no NCD at baseline and risk of cognitive impairments at 18 and 36-months follow-up

Longitudinal LMEs revealed no significant relationship between baseline BAG and NCD at 18-and 36 months in those with no NCD at baseline. In contrast, survival analysis among the same initially non-impaired patients revealed that the patients belonging to the upper BAG quartiles at baseline were more likely to develop NCD during the follow-up period, indicating that a younger appearing brain represents a protective factor for longitudinal cognitive decline after stroke.

Among the 129 patients with no NCD at baseline, 24 showed evidence of cognitive decline at 18 months and 12 at 36 months. Although the survival analysis supported the clinical relevance of BAG for long-term prognosis, the lack of predictive value of acute-phase BAG for long-term decline suggested by the LMEs may be explained by non-random study attrition at the 36 months assessment and low power at the follow-up examinations. Further, while our study design does not allow us to test this hypothesis, it is conceivable that pre-stroke BAG is a more reliable predictor for future decline than post-stroke BAG. More studies on the use of early-phase BAG as a predictor for future risk of cognitive impairments in those with no initial NCD is needed.

Brain aging is associated with a natural ‘wear and tear’, but is highly influenced by environmental factors throughout life. Although linked to genetics (Kaufmann et al., 2019; Cole et al., 2017) and birth weight (Vidal-Pineiro et al., 2021), brain structure and related brain age is associated with a range of proxies for general and cardiometabolic health (Montine et al., 2019; Beck et al., 2022a; Beck et al., 2022b) and life-style choices such as unhealthy diet (Onaolapo et al., 2019), lack of physical exercise (Stillmann et al., 2020; Sanders et al., 2021) and heavy alcohol use (Sullivan & Pfefferbaum, 2019). In line with our current findings, a younger appearing brain is not only associated with lower risk of cognitive deficits, but also increases resilience to consequences after injury, such as a stroke (Montine et al., 2019).

### 4.3 Preserved cognitive function across the follow-up period and brain changes over time

Our analysis comparing rate of BAG change with the patients showing preserved cognitive status revealed no significant group difference. The increasing BAG over time in the declining group indicates a connection between BAG and cognition over time, which emphasizes the link between preserved brain structure and cognitive functions. The current study is the first (to the author’s knowledge) one of its kind, looking at the link between BAG and the preservation of cognition after stroke. Findings from non-stroke cohorts do however exist. One study looking at BAG and NCD in non-stroke participants found overall higher BAG in participants exhibiting progressive mild NCD compared to those with a preserved mild NCD over time, although this difference was not statistically significant (Popescu et al., 2021). Also, a preserved cognitive function in processing speed over the course of 2.5 years in functionally intact older adults has been associated with larger corpus callosum volume and lower levels of inflammation (Bott et al., 2017). Such superior agers exhibit similar memory ability as participants 20-30 years younger and also show less cortical atrophy compared to younger controls (Harrison et al., 2012), and less atrophy of the hippocampus and other regions of the default mode and salience networks (Sun et al, 2016) compared to age-matched controls. Postmortem studies also find less Alzheimer type neurofibrillary tangles in these superior agers (Gefen et al., 2015) and they have a reduced risk for cognitive impairment (Dang et al., 2019). This, taken together with our findings, signifies the value of using brain age rather than chronological age in post stroke NCD prediction.

### 4.4 Strengths and limitations

This study has some major strengths, such as utilizing a large dataset that incorporates MRI, clinical and neuropsychological examinations over the course of three years following a stroke. The longitudinal perspective enables the identification of factors associated with preserved cognition and delays in the onset of post stroke cognitive decline. While higher clinical and investigator variability related to the multi-center design may reduce statistical power, the naturalistic multi-study design increases generalizability.

This study also has limitations that need consideration. First, although brain age prediction is based on clinically available MRI data, the analysis still requires a substantial amount of processing. To increase feasibility in a clinical context future work may implement novel approaches allowing for accurate estimations based on e.g. deep neural networks and minimally processed MRI data (Leonardsen et al., 2021). Next, study attrition at 36 months was considerable, leading to reduced statistical power and possible biases. Attrition is a typical challenge in longitudinal studies, especially when investigating disease in older participants (Teague et al., 2018). Participants who remained in the study throughout the follow-up period were generally healthier and had better cognition than those lost to follow-up (supplementary figure 6). Lastly, we primarily included patients with mild to moderate strokes, reducing generalizability to patients suffering severe strokes. In comparison to the Norwegian Stroke Registry, Nor-COAST patients have previously been found to be representative of the majority of the Norwegian stroke population, primarily suffering from mild strokes (Kuvås et al., 2020). Automated software for segmentation and image reconstruction in brains with more severe strokes is challenging due to the lesion itself, but also severe strokes making it more challenging for the patient to undergo MRI (Ozzoude et al., 2020). Future work could prevent selection bias through utilizing standard clinical protocols rather than requiring a second (study-specific) MRI at baseline.

## Conclusion

Our analyses revealed that higher BAG was associated with post stroke cognitive impairment across the follow-up time, indicating that a younger appearing brain increases resilience and is associated with lower risk for post stroke NCD up to 36 months after a stroke. The current study indicate that this may also be true for those with no cognitive impairment 3 months after stroke. The findings show that BAG is sensitive to cognitive impairment after stroke and may be used in order to predict cognitive outcome in stroke patients.

## Data Availability

All data produced in the present study are available upon reasonable request to the authors

## Supplementary material

**Supplementary table 1:** Linear mixed model results per analysis

|  | ANALYSIS I |  |  | ANALYSIS II |  |  | ANALYSIS III |  |  |
| --- | --- | --- | --- | --- | --- | --- | --- | --- | --- |
|  | Coef. | CI | p | Coef. | CI | p | Coef. | CI | p |
| **NCD** | **1.37** | **.33 to 2.42** | **.010** | .56 | -1.05 to 2.18 | .50 | .27 | -1.11 to 1.64 | .71 |
| **Time** | -.30 | -.79 to .19 | .23 | -.57 | -1.25 to .10 | .095 | .04 | -.77 to .85 | .92 |
| **NCD#time** | .50 | -.51 to 1.52 | .33 | .71 | -2.34 to 3.76 | .65 | -.78 | -3.68 to 2.12 | .60 |
| **Age** | -.03 | -.12 to .05 | .47 | -.05 | -.16 to .06 | .35 | -.05 | -.17 to .08 | .44 |

**Supplementary table 2:** Estimated margins of NCD status and time interactions across analyses

|  | ANALYSIS I |  | ANALYSIS II |  | ANALYSIS III |  |
| --- | --- | --- | --- | --- | --- | --- |
|  | **Margins** | **CI** | **Margins** | **CI** | **Margins** | **CI** |
| **No-NCD** (“preserved” in A-III) # **Baseline** | -.17 | -1.37 to 1.03 | -1.54 | -2.91 to -.16 | -.74 | -2.55 to 1.08 |
| **Any-NCD** (“changed” in A-III) # **Baseline** | .65 | -.56 to 1.87 | -2.16 | -6.67 to 2.35 | .59 | -3.78 to 4.95 |
| **No-NCD** (“preserved” in A-III) # **18 mo.** | -.68 | -1.81 to .44 | -2.14 | -3.49 to -.78 | -.57 | -1.96 to .82 |
| **Any-NCD** (“changed” in A-III) # **18 mo.** | .64 | -.48 to 1.76 | -2.05 | -4.24 to .14 | -.02 | -2.30 to 2.25 |
| **No-NCD** (“preserved” in A-III) # **36 mo.** | -1.20 | -2.59 to .19 | -2.74 | -4.39 to -1.10 | -.40 | -1.92 to 1.12 |
| **Any-NCD** (“changed” in A-III) # **36 mo.** | .63 | -.85 to 2.11 | -1.95 | -4.64 to .75 | -.64 | -2.72 to 1.44 |

**Supplementary table 3:**
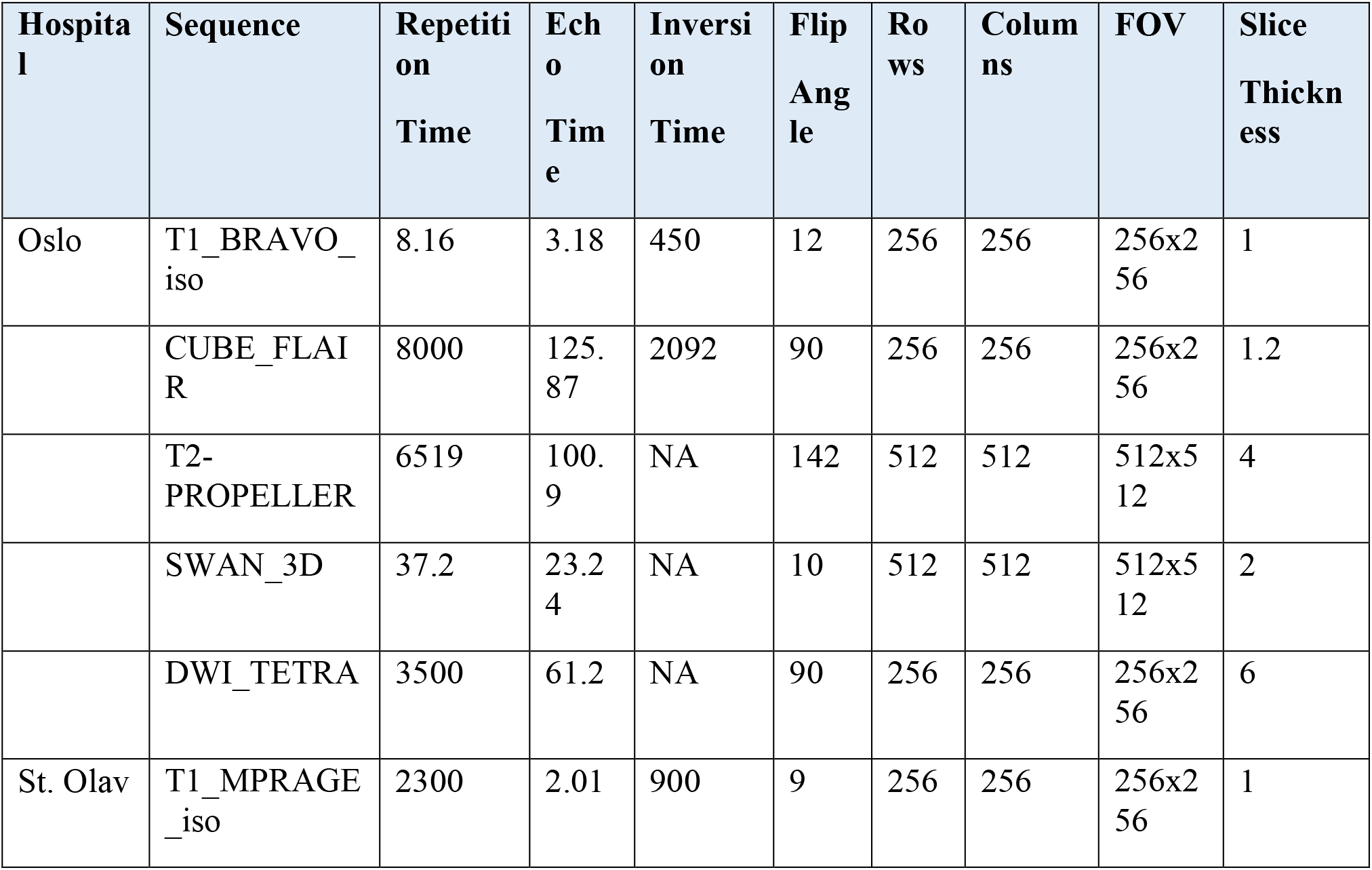
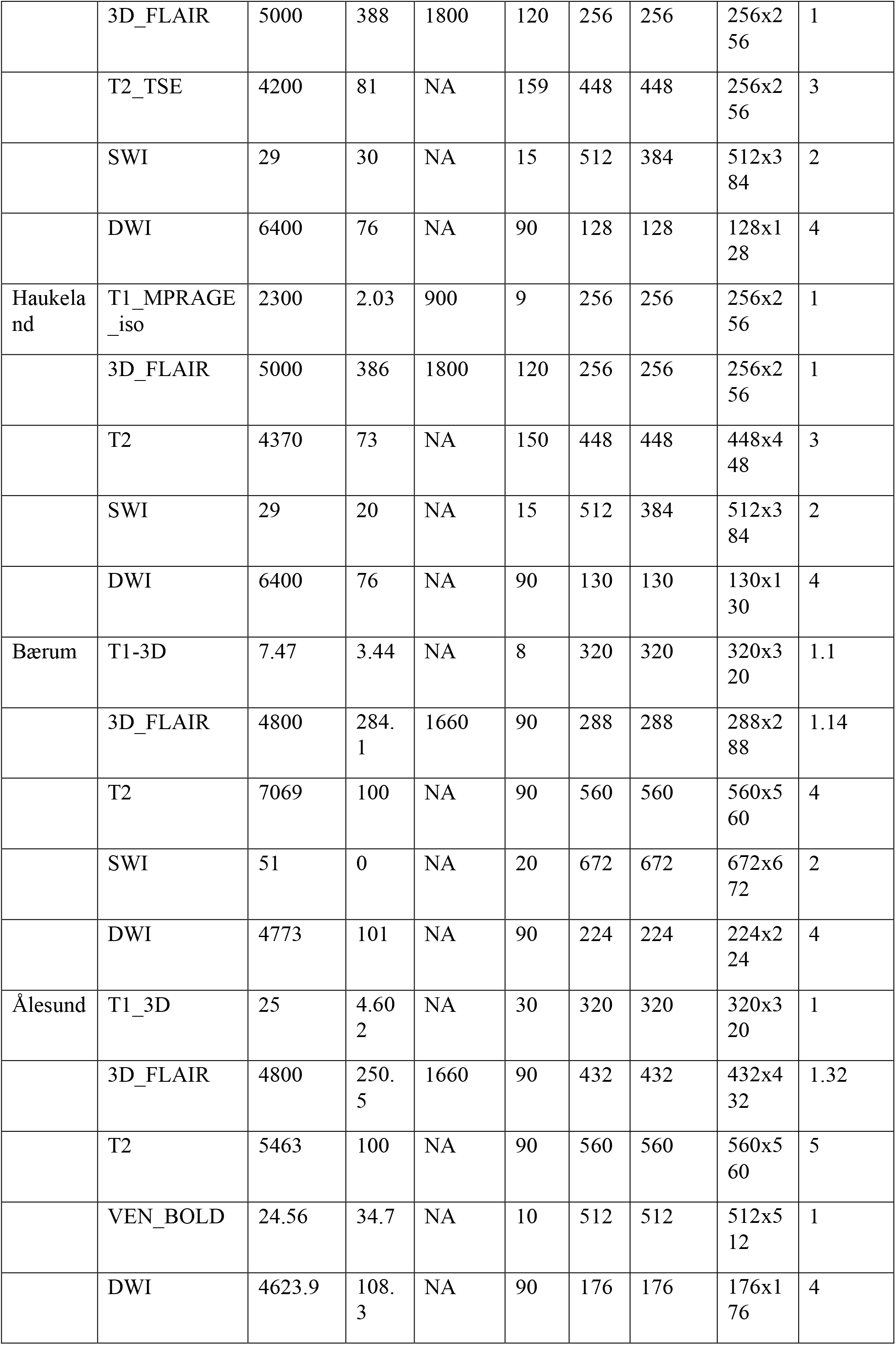
List of MRI sequence parameters across hospitals

| **Hospital** | **Sequence** | **Repetition Time** | **Echo Time** | **Inversion Time** | **Flip Angle** | **Rows** | **Columns** | **FOV** | **Slice Thickness** |
| --- | --- | --- | --- | --- | --- | --- | --- | --- | --- |
| Oslo | T1_BRAVO_iso | 8.16 | 3.18 | 450 | 12 | 256 | 256 | 256x256 | 1 |
|  | CUBE_FLAIR | 8000 | 125.87 | 2092 | 90 | 256 | 256 | 256x256 | 1.2 |
|  | T2-PROPELLER | 6519 | 100.9 | NA | 142 | 512 | 512 | 512x512 | 4 |
|  | SWAN_3D | 37.2 | 23.24 | NA | 10 | 512 | 512 | 512x512 | 2 |
|  | DWI_TETRA | 3500 | 61.2 | NA | 90 | 256 | 256 | 256x256 | 6 |
| St. Olav | T1_MPRAGE_iso | 2300 | 2.01 | 900 | 9 | 256 | 256 | 256x256 | 1 |
|  | 3D_FLAIR | 5000 | 388 | 1800 | 120 | 256 | 256 | 256x256 | 1 |
|  | T2_TSE | 4200 | 81 | NA | 159 | 448 | 448 | 256x256 | 3 |
|  | SWI | 29 | 30 | NA | 15 | 512 | 384 | 512x384 | 2 |
|  | DWI | 6400 | 76 | NA | 90 | 128 | 128 | 128x128 | 4 |
| Haukeland | T1_MPRAGE_iso | 2300 | 2.03 | 900 | 9 | 256 | 256 | 256x256 | 1 |
|  | 3D_FLAIR | 5000 | 386 | 1800 | 120 | 256 | 256 | 256x256 | 1 |
|  | T2 | 4370 | 73 | NA | 150 | 448 | 448 | 448x448 | 3 |
|  | SWI | 29 | 20 | NA | 15 | 512 | 384 | 512x384 | 2 |
|  | DWI | 6400 | 76 | NA | 90 | 130 | 130 | 130x130 | 4 |
| Bærum | T1-3D | 7.47 | 3.44 | NA | 8 | 320 | 320 | 320x320 | 1.1 |
|  | 3D_FLAIR | 4800 | 284.1 | 1660 | 90 | 288 | 288 | 288x288 | 1.14 |
|  | T2 | 7069 | 100 | NA | 90 | 560 | 560 | 560x560 | 4 |
|  | SWI | 51 | 0 | NA | 20 | 672 | 672 | 672x672 | 2 |
|  | DWI | 4773 | 101 | NA | 90 | 224 | 224 | 224x224 | 4 |
| Ålesund | T1_3D | 25 | 4.602 | NA | 30 | 320 | 320 | 320x320 | 1 |
|  | 3D_FLAIR | 4800 | 250.5 | 1660 | 90 | 432 | 432 | 432x432 | 1.32 |
|  | T2 | 5463 | 100 | NA | 90 | 560 | 560 | 560x560 | 5 |
|  | VEN_BOLD | 24.56 | 34.7 | NA | 10 | 512 | 512 | 512x512 | 1 |
|  | DWI | 4623.9 | 108.3 | NA | 90 | 176 | 176 | 176x176 | 4 |

**Supplementary table 4:** Hospital MRI, BAG and NCD-status numbers

| Hospital |  | Baseline | 18 months | 36 months |
| --- | --- | --- | --- | --- |
| Ullevål | MRI | 48 | 43 | - |
|  | BAG | 33 | 32 | - |
|  | NCD-status | 28 | 32 | - |
| Bærum | MRI | 60 | 55 | 37 |
|  | BAG | 63 | 56 | 35 |
|  | NCD-status | 62 | 50 | 30 |
| Haukeland | MRI | 43 | 33 | - |
|  | BAG | 43 | 32 | - |
|  | NCD-status | 35 | 29 | - |
| St Olav | MRI | 193 | 106 | 74 |
|  | BAG | 167 | 95 | 69 |
|  | NCD-status | 138 | 91 | 66 |
| Ålesund | MRI | 8 | 4 | - |
|  | BAG | 8 | 4 | - |
|  | NCD-status | 6 | 4 | - |
| Sum | MRI | 352 | 241 | 111 |
|  | BAG | 314 | 219 | 104 |
|  | NCD-status | 269 | 206 | 96 |

**Supplementary table 5:**
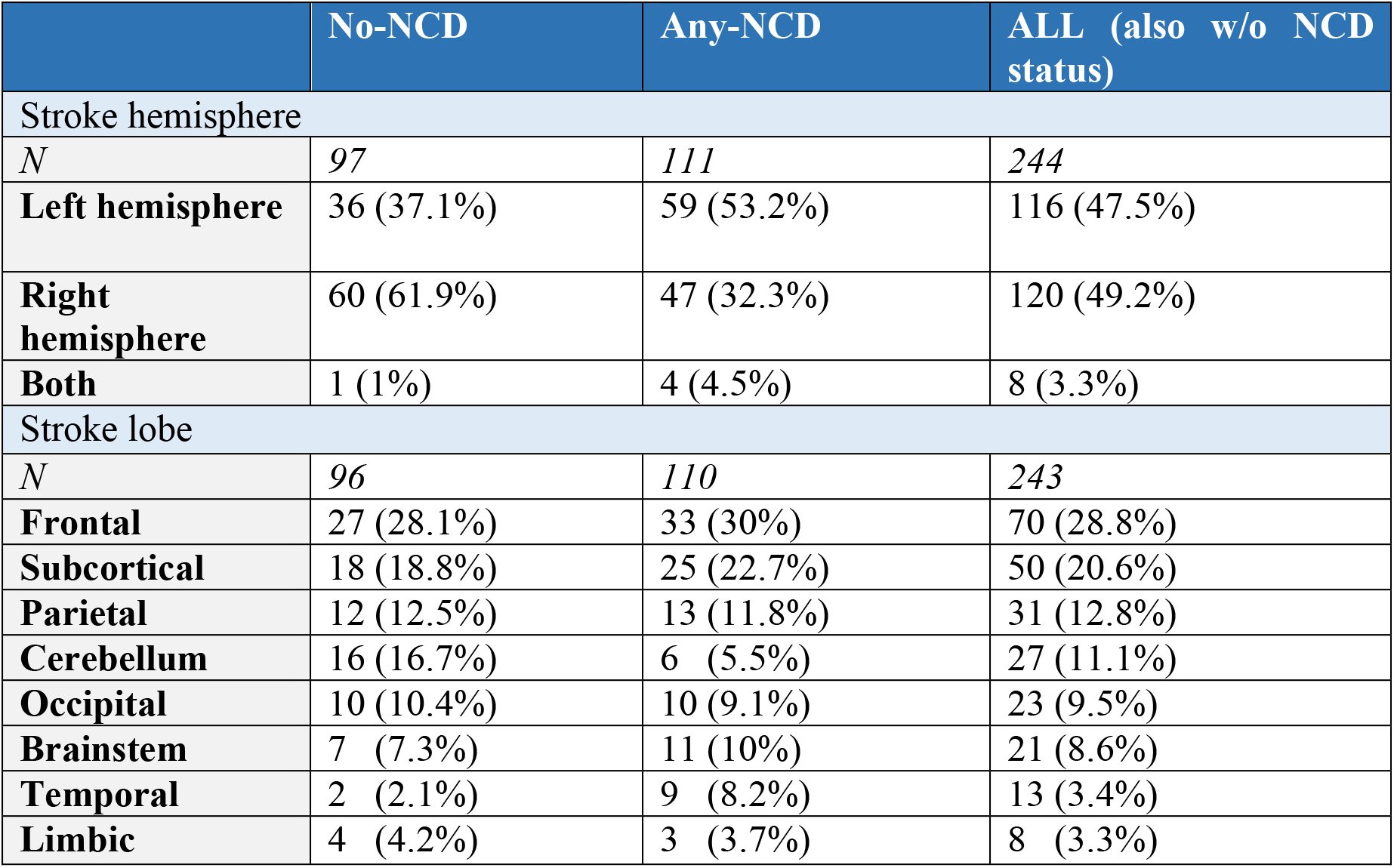
Stroke location by NCD status

**Supplementary table 6:** Characteristics of participants with 1, 2 or 3 timepoints

|  | ONLY 1 TIMEPOINT | 2 TIMEPOINTS | 3 TIMEPOINTS |
| --- | --- | --- | --- |
| *N* | 163 | 201 | 72 |
| Women | 82 (50.3%) | 116 (57.7%) | 42 (58.3%) |
| Age | 73.2 (12.6) | 70.4 (10.8) | 71.1 (8.5) |
| Education (years) | 12.3 (3.9) | 12.6 (3.7) | 12.5 (3.9) |
| GDS (pre-stroke) | 1.6 (1.0) | 1.3 (.6) | 1.2 (.4) |
| mRS | 2.3 (1.4) | 1.9 (1.2) | 1.8 (1.0) |
| NIHSS | 4.3 (4.9) | 3.7 (4.9) | 4.1 (5.9) |
| MoCA | 23.6 (5.3) | 25.3 (3.8) | 26.3 (3.0) |
| NCD normal | 47 (28.8%) | 105 (52.2%) | 42 (58.3%) |
| NCD mild | 29 (17.8%) | 61 (30.4%) | 19 (26.4%) |
| NCD major | 45 (27.6%) | 29 (14.4%) | 10 (13.9%) |
| Stroke size (mL) (N) | 15.9 (31.9) | 9.4 (26.4) | 8.1 (18.6) |
| Med temp thick (mm) (N) | 2.82 (.16) | 2.87 (.17) | 2.97 (.11) |

## Notes

### Competing Interest Statement

The authors have declared no competing interest.

### Funding Statement

The study was funded by a research grant from the South-Eastern Norway Regional Health Authority (Helse Sor-Ost).

### Author Declarations

Nor-COAST was approved by the regional committee for medical and health research, REK Nord (REK number: 2015/171), and registered on clinicaltrials.gov (NCT02650531). REK Nord has also approved this current sub study (REK number: 2019/397).

